# Can Public Libraries Be Leveraged to Expand Access to Telehealth? Exploration of a Strategy to Mitigate Rural Health Disparities

**DOI:** 10.1101/2020.08.18.20177287

**Authors:** Pamela B. DeGuzman, Neha Jain

## Abstract

In the U.S., those who lack broadband internet have limited ability to connect to care providers over a telemedicine video visit (VV). During the coronavirus disease pandemic, VVs have become increasingly common, but are not equitably accessible, which may exacerbate existing health disparities. Widening health disparities are of particular concern in the rural U.S. where broadband is lacking. We term this inequity in healthcare access due to limited internet access the “digital health divide.” Because public libraries typically offer free use of broadband internet to patrons, they can help bridge the digital health divide and assist patrons with VVs. However, no guidelines currently exist for care providers and libraries to implement this needed, but potentially complex undertaking. Individual programs in which community members have used public libraries as a place from which to connect to a VV may offer insight into guidance needed. Thus, we conducted a scoping review to explore interventions reporting use of public libraries for community members to connect to a healthcare provider via telemedicine. One article was found describing the use of a public library for community members to connect to a telemedicine VV. The use of public libraries as spaces from which patrons can participate in VVs with providers is promising, but research is urgently needed to guide implementation.

---

In the United States, there is clear evidence that rural populations experience significant health disparities compared with their urban counterparts. Rural U.S. residents are more likely to have chronic diseases such as hypertension, have worse health outcomes, and are less likely to participate in a variety of health-promoting behaviors (Matthews et al. 2017; Douthit et al. 2015). Residents of rural areas are more likely to die from cancer, despite lower overall cancer incidence rates (Henley et al. 2017). Diabetes and heart disease are not only far more prevalent among rural Americans than their urban counterparts, but also associated with a higher mortality rate (Callaghan et al. 2019; O’Connor and Wellenius 2012). Higher rates of certain cancers and other chronic illness are likely related to the lower rates of health promoting behaviors among rural Americans; those from rural areas are more likely to smoke and less likely to maintain an optimal body weight and meet physical activity recommendations than their urban counterparts (Douthit et al. 2015). Although overall cancer prevalence is lower in rural areas, the cancers associated with modifiable behaviors (smoking, diet, physical activity) are higher (Zahnd et al. 2018). Mental health outcomes are also less favorable to those living in rural areas: suicide rates are consistently higher among rural populations than those living in more urbanized areas, and among younger rural populations rates are nearly twice as high as their urban counterparts (Fontanella et al. 2015; Ivey-Stephenson et al. 2017).

Disparities in disease screening, health education, and modifiable behaviors between rural and urban populations are in large part related to the scarcity of healthcare prevention and treatment services available in rural areas (Nelson et al. 2020). For example, residents of rural American have are less likely to be receive screening for cancer and cardiovascular diseases (Caldwell et al. 2016; Schroen and Lohr 2009), and despite higher rates of diabetes, over half of all rural counties in the U.S. do not have a diabetes education management program (Centers for Disease Control and Prevention 2018).

Overcoming these disparities must include improving *access to care*, which includes the “timely use of personal health services to achieve the best health outcomes” (RUPRI Health Panel 2014). Unfortunately, the recent 2019 coronavirus disease (COVID-19) pandemic has likely widened disparities in access to care. In recent months, healthcare providers have increased their use of telemedicine systems to provide video visits (VV), via which providers can visually connect with patients while allowing both parties to minimize disease exposure (Mann et al. 2020). There have been multiple calls to use telehealth to improve access to rural populations across health and disease states (Marcin, Shaikh, and Steinhorn 2016; Douthit et al. 2015; Zahnd and Ganai 2019). However, because VVs require users to have access to a fixed broadband signal of sufficient strength to stream a video call, the inequitable distribution of broadband in the U.S. creates disparities in access to this technology, most notably in rural areas (DeGuzman et al. 2020).

The difference in easy access to fixed broadband between groups is commonly referred to as the *digital divide* (Bertot, Real, and Jaeger 2016). In the U.S., fixed broadband is less available in rural areas, and when it is available, it is often cost-prohibitive as to be effectively unavailable (Federal Communication Commission 2019; LaRose et al. 2007). *Digital inclusion*, which refers to not only the access to, but also the skills to use the internet, further compounds the digital divide (Rhinesmith 2016). In rural areas, those lacking experience using the Internet and broadband are less likely to use Internet technology, leading to difficulty utilizing digital technology to improve health access (LaRose et al. 2007; DeGuzman et al. 2020). In this paper, we refer to the inability of community members to access a VV due to either lack of a broadband signal or limited digital inclusion as the *digital health divide* (DHD).

To help mitigate the DHD, rural health disparities researchers and advocates have suggested use of public libraries as a place from which rural populations lacking home-based broadband can connect to a VV (“A National Public Policy Agenda for Libraries and the Policy Revolution Initiative” 2015; Clapp 2010; DeGuzman, Siegfried, and Leimkuhler 2020). In fact, libraries may be an optimal solution: Virtually all public libraries offer publicly available internet access (Real and Rose 2017). Public librarians are highly trained in information access skills, which can help bridge the digital inclusion gap (Bertot, Real, and Jaeger 2016), and they may regularly assist patrons with accessing health information (Strover 2019). Public libraries are frequented by older adults seeking information who are less likely to regularly access the internet, and more likely to require health care services (DeGuzman, Siegfried, and Leimkuhler 2020; Levy, Janke, and Langa 2015). In fact, conducting a visit in a public library may even help overcome psychological barriers that rural populations encounter with seeking health care services(Douthit et al. 2015): Rural residents have acknowledged that using telehealth services in a public library is a reasonable option because of the lack of stigma associated with being seen visiting a library (Sundstrom et al. 2019).

For both health care providers and public librarians, the decision to collaborate on an achievable strategy for community members to access a VV may be obvious; however, the implementation of such an undertaking is complex. Private space, internet speed, and equipment available at the library all need to be considered, as well as receptivity of both patient and library staff, and potential scheduling barriers (DeGuzman, Siegfried, and Leimkuhler 2020). Guidelines for initializing and implementing a partnership between libraries and providers may help improve adoption of this practice in communities where residents have limited broadband access. A first step towards developing guidelines is to gain enhanced understanding of how current interventions have navigated these complexities. Thus, we conducted a review of literature to explore and map relevant literature. Given the cross-disciplinary nature as well as the need to examine the range of research activity, we selected a scoping methodology to explore and describe any research in which public libraries were used as sites for community members to connect to telemedicine (Arksey and O’Malley 2005).

## Method

We followed the framework of Arksey and O’Malley to identify and map literature. Their scoping framework directs reviewers through 5 stages: *Identifying the research question, identifying relevant studies, study selection, charting the data and collating, summarizing and reporting the results*.

### Identifying the Research Question

The first step in the literature scoping method is to identify the research question, including the relevant study population, and any interventions or outcomes of interest. The research question that guided our review was *What is known from existing literature about interventions that report or evaluate community members connecting to a healthcare provider using telemedicine in a community public library?*

### Identifying Relevant Studies

Arksey and O’Malley’s suggest casting a wide net to find literature from various disciplines. We opted to search multiple electronic databases across both library science and health, and amended our search reference lists of review articles, and a hand search of key journals. With guidance from a health librarian at the University of Virginia Health Sciences Library, we searched five databases from both health and library disciplines to find articles to include in the review: PubMed, the Cumulative Index for Nursing and Allied Health Literature; Web of Science (WOS); Library, Information, Science & Technology Abstracts; and Library Literature Full Text. In each database, we searched for Medical Subheading (MeSH) terms “Telemedicine” OR “Telehealth” AND “Libraries,” except for in WOS we searched for these terms using a full text search, rather than MeSH terms, and limited to *article* document types. All searches were restricted to English language and between 2005 and 2020. All database searching was conducted in June 2020. We further searched the reference list of review articles that were discovered through electronic database search, searching for articles with “library” or “libraries in the title. We searched two journals: *Public Libraries Quarterly* and *Health Information and Libraries Journal* using the online search feature provided by the publisher of each journal. Both journals were searched for terms “telemedicine” and “telehealth.”

### Study Selection

Study selection for the articles identified in the electronic database search was guided by the Preferred Reporting Items for Systematic Reviews and Meta-Analyses (PRISMA) statement (Moher et al. 2009). All duplicates were identified using the *Check for Duplicates* tool in Mendeley (Elsevier; Amsterdam, the Netherlands), and removed. We screened these articles using two inclusion criteria. To be included, articles had to (1) describe a telemedicine intervention in which patients were connected with any type of healthcare provider, and (2) be conducted with patients in a public (i.e. not hospital or university) library. Initially, one researcher (N.J.) screened titles and abstracts of all articles to assess for inclusion criteria. To ensure reliability of screening results, a second researcher (P.B.D) screened a random sample of 10% of all reviewed articles to assess agreement with exclusions. For those not excluded on the basis of titles and abstracts, full texts were reviewed and discussed by the two researchers to determine if they fit inclusion criteria. We conducted a similar screening for articles identified through our search of review article reference lists and key journal searches. First all duplicates were removed, then any remaining articles were screened using the inclusion criteria.

### Charting the Data and Summarizing the Results

After both levels of screening, a narrative review was conducted to chart and summarize all remaining articles to identify key aspects of each intervention implementation. Specifically, we evaluated each article with regard to ensuring privacy, adequacy of Internet speed, and equipment requirements, receptivity of both patient and library staff, and potential scheduling barriers.

## Results

Results of article identification and screening are presented in a flow diagram (see Figure). We identified 258 articles between the five databases, 42 of which were review articles. We identified an additional 14 articles through the key journals search and one additional article through a hand search. No articles were found through a search of the reference lists of the reviews. After 29 duplicates were removed, 244 articles remained. Through title and abstract review, we determined that 225 did not meet inclusion criteria. Agreement between both researchers of articles excluded at this step was 100%. Both researchers reviewed the full-text of the remaining six articles; five were excluded because interventions were conducted in either a hospital or university library, not a public library. One article remained for analysis.

**Figure 1:**
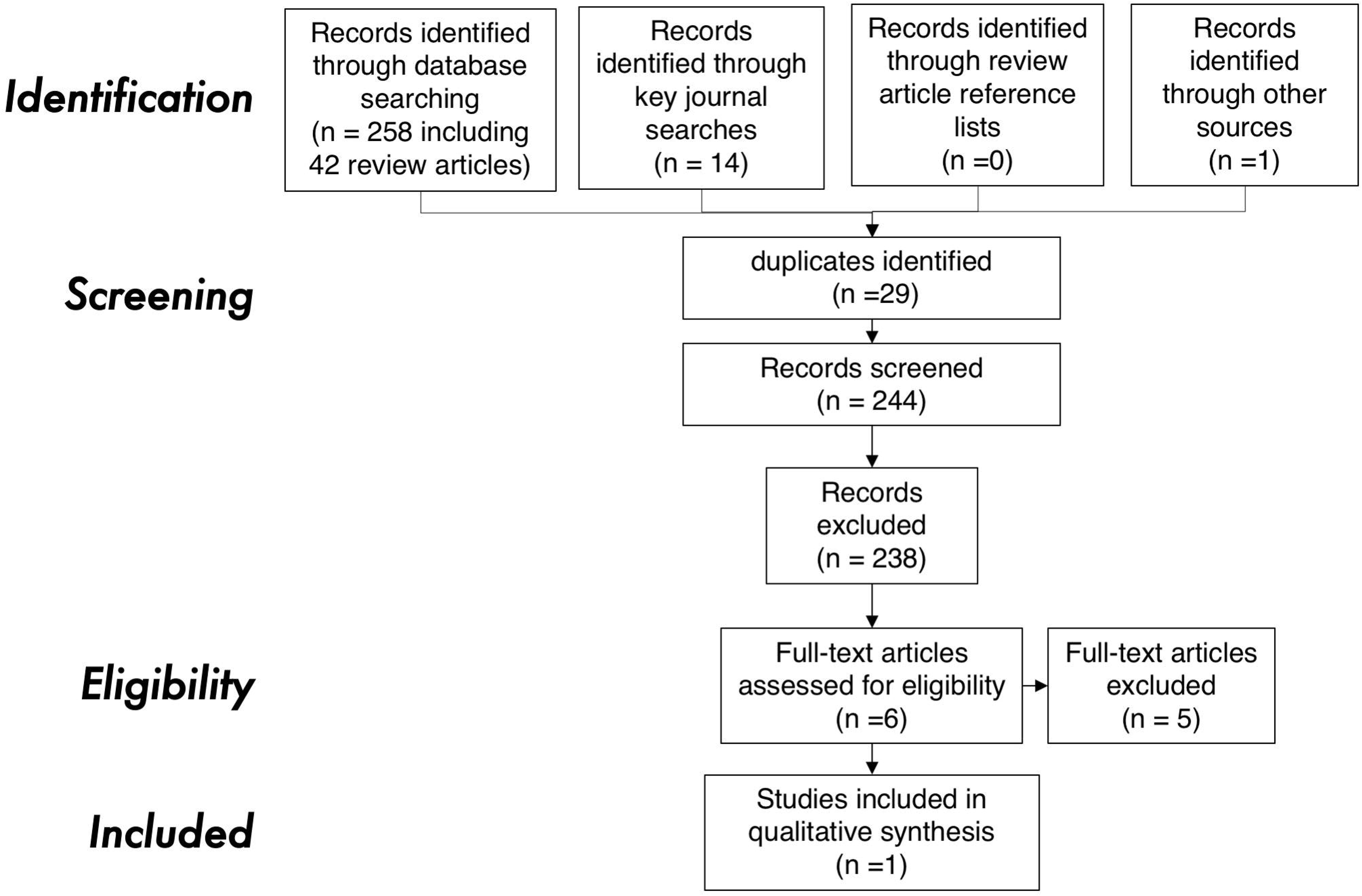
Identification, screening, eligibility, and inclusion of articles. Adapted from the Preferred Reporting Items for Systematic Reviews and Meta-analyses flow diagram (http://www.prisma-statement.org/PRISMAStatement/FlowDiagram).

In 2011, McIlhenny and colleagues published a description of their telemedicine-delivered educational intervention for patients with Type 2 diabetes who received care at one of two rural clinics. Participants could access the intervention from their homes if they had broadband internet; otherwise they were instructed in alternative methods of connecting to the intervention: they could use their own or a borrowed laptop at either one of the two clinics, and were also “informed of public venues where they could access the internet (e.g. public library) if they desired.” (McIlhenny et al. 2011)^(p.4)^. In the reporting of the study, the researchers did not appear to coordinate with specific libraries, and no data about libraries’ available privacy, hardware or broadband capabilities, receptivity of patient and library staff, or potential scheduling barriers were discussed.

## Discussion

Our scoping review revealed one article in which a telemedicine intervention was intended by researchers to be conducted from a public library. It is possible that other interventions have been conducted, but have not been reported in the peer-reviewed literature. The researchers did not report how many participants used the library to connect to the intervention, nor did they describe equipment or resources available at libraries (McIlhenny et al. 2011).

Our analysis of current literature was limited by searching only five databases, and having only two individuals search the literature. It is possible that casting a wider net with a larger research team could uncover additional research that has evaluated this practice. It is also possible that no libraries or care providers have implemented this type of program. Although public libraries are a major hub for internet use in rural areas, the individual nature of rural public libraries’ administrative structures make coordinating services with libraries a complex undertaking (Strover 2019). Whereas large public library systems have close to 100 branches (New York Public Library 2020) with operating budgets in excess of $5 million (American Library Association 2020b), rural public libraries are typically one small, single unit. In fact, over 60% of rural libraries are a single-library system (Real and Rose 2017). Thus, any care provider --from large health systems providing regional specialty care to individual community-based providers--must investigate and negotiate with each individual library to ensure adequate space, equipment, and broadband speed availability, adding to the challenge of connecting with rural patients.

Our research did not yield information that could be used to systematically guide library-practice partnerships to develop telemedicine hubs in public libraries. In the context of COVID-19 restrictions both increasing the use of VV and the increased risk of disease exposure when visiting a practice site, research is urgently needed to help develop these hubs in rural libraries. Understanding both librarians’ and healthcare providers’ perspectives on implementation considerations can help speed development and testing of interventions to help bridge the DHD in rural communities.

### Conclusions

COVID-19 has increased the DHD, placing those without broadband access at further disadvantage. As rural health disparities continue to widen, novel solutions are urgently needed to safely connect community members with care providers to ensure equitable access to care. Many types of prevention-based health care can be delivered via a telemedicine VV, such as counseling and education. As public libraries around the country begin to re-open in response to COVID-19 restriction easements, librarians anticipate that 60% of patrons will return to libraries in search of electronic resources (American Library Association 2020a). The use of public libraries are places from which patrons can participate in a VV with their providers is promising, but we lack research guiding these potentially complex partnerships. Given the increasing awareness of the digital divide in the U.S., now is the time to act.

## Data Availability

all data is available through library databases

## Notes

The authors have no conflicts of interest to declare.

### Competing Interest Statement

The authors have declared no competing interest.

### Funding Statement

no funding was received for this work

### Author Declarations

No human subjects were involved in this work

